# Synthetic Validation of Pediatric Trust Instruments using Persona-Driven Large Language Models

**DOI:** 10.1101/2025.11.25.25340922

**Authors:** Ella Boone, Katya Loban, Elena Guadagno, Dan Poenaru

## Abstract

**Objectives:** Trust is foundational to patient-physician relationships and is associated with improved care-seeking and adherence in primary care. However, validated trust instruments for pediatric emergency and surgical contexts are lacking, and traditional instrument development is slow and resource-intensive. Large language models (LLMs) could streamline the validation process by serving as scalable, systematic expert panel surrogates.

**Materials and Methods:** We developed four new trust assessment instruments: two for patient-families and two for physicians. Two-phase content validation was conducted using two parallel synthetic and human expert panels. Synthetic panels consisted of three persona-prompted LLMs (Claude Sonnet 4, GPT-5, Grok4). Human panels served as traditional comparators. Scale-Content Validity Index (S-CVI) and Fleiss’ kappa (κ) acceptance thresholds were set at ≥0.80.

**Results:** Combined human–synthetic expert panels revealed substantial inter-rater reliability across all instruments. Fleiss’ κvalues for dimensional validation were: patient-family = 0.84 (95% CI [0.72, 0.96]), physician = 0.87 (95% CI [0.72, 1.00]);contextual validation: patient-family = 0.83 (95% CI [0.73, 0.93]), physician = 0.88 (95% CI [0.80, 0.96]). All instruments exceeded S-CVI ≥0.80 thresholds across both validation phases.

**Discussion:** Persona-prompted LLMs demonstrated comparable validity outcomes to human experts while accelerating validation timelines from months to weeks. Future research needs to evaluate this approach across psychometric testing phases.

**Conclusion:** This synthetic instrument validation methodology offers a scalable blueprint for healthcare measurement development, enabling faster creation of validated tools to support evidence-based patient care.

## BACKGROUND

Trust forms the foundation of patient-physician therapeutic relationships. We define trust in this study as the patient’s acceptance of a vulnerable situation and their confidence that the physician will care for them.[1,2] Patient trust is prospective, complex, and based on expectations of care from a provider.[3] The experience of trust is well-established in primary care settings, with increased trust associated with greater care satisfaction, treatment adherence, and continuity of care.[4–6] Trust can also facilitate care seeking by increasing patient willingness and disclosure of information during clinical encounters.[7,8]

Trust in brief, inherently stressful and complex medical settings such as emergency and surgical care, requires different relationships, and therefore distinct conceptualization.[9–12] Studies have examined modified scales developed for adult primary care settings in pediatric emergency and surgical care, yet none have evaluated the nuances of trust-building in these unique settings.[1,9,13] In a recently published scoping review our team has identified a gap in the literature regarding how trust is built between patient-families and physicians in pediatric surgical and emergency settings.[1] Therefore, this study aims to design an instrument to measure trust and its key components in these settings, from both patient-family and physician perspectives.

Designing new healthcare instruments is a complex, time- and resource-intensive process. This can take months to years, and many new instruments therefore remain undeveloped, unable to impact patient care.[14,15] When these time and resource constraints are considered alongside the recruitment challenges associated with engaging content experts and families within the specialized field of pediatric urgent and surgical care, it is likely that the development of our intended instrument will encounter similar barriers.[16]

Artificial Intelligence (AI) is rapidly changing how resources are utilized in healthcare.[17] Synthetic AI data have become increasingly acceptable for validating text classification, data-driven decision-making, and predictive analytics.[18,19] We hypothesize that such synthetic data can address the barriers of traditional expert panel recruitment and reduce instrument validation time. Generating synthetic data for content validation requires specific ‘expert’ roles to provide item ratings, by using large language models (LLMs) through persona prompting.[20,21]

This study aims to develop a method for preliminary content validation of healthcare survey instruments using a synthetically generated expert panel in parallel to a smaller traditional human expert panel. This methodological approach, in which we term *synthetic instrument validation* (SIV), positions LLMs as complementary to human experts, not as replacements, highlighting the synergistic relationship between human and AI capabilities.[22] We expect that this approach will reduce time and recruitment constraints, and facilitate instrument development for data-informed patient care improvements in pediatrics and beyond.

## METHODS

We developed and conducted a preliminary validation of four instruments to assess trust levels and its components by measuring four dimensions of trust: competence, caring, communication, and dependability (Figure 1).[1]

**Figure 1.**
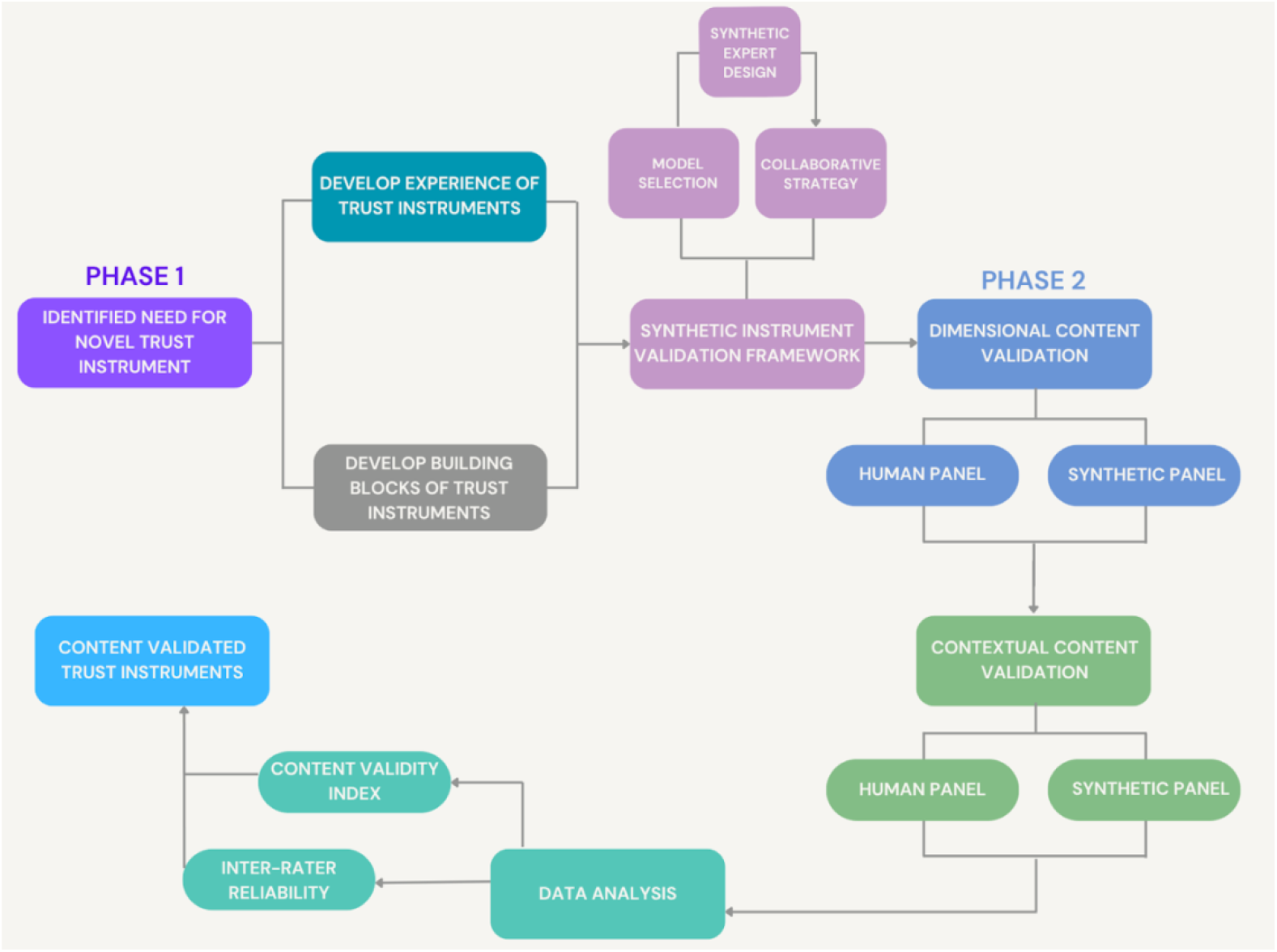
Overview of study methods for trust instrument development and validation

### Instrument Development

### Phase 1: Instrument Design

#### Adaptation of Trust Instruments

Elements from Hall et al.’s *Trust in Physician Scale*[23] were adapted for pediatric surgical and emergency contexts as outlined in **Supplementary Material S1**. The 5-point Likert scale was modified to a 4-point (1 = No, not at all; 4 = Yes, definitely) by removing the neutral midpoint. Verb tense was modified to emphasize acute rather than longitudinal care, and selected items used first-person phrasing (“I felt”) to capture patient-family perspective. Eight of 10 items best fitting the study context were selected and four new items were added. This patient-family instrument was named the *Trust-Experience-Patient-Family* (TRUST-E-F).

Review panel meetings with a pediatric general surgeon, general surgery resident, two pediatric emergency physicians, and a quantitative researcher organized items into the four trust dimensions. Wording was modified to align each item with a single dimension,per COnsensus-based Standards for the Selection of healthcare Measurement INstruments (COSMIN) guidelines.[24]

The physician instrument (*Trust-Perception-Physician*, TRUST-P-MD) corresponded directly to ETP items but was framed from the physician perspective using the same 4-point Likert scale. Similar expert meetings classified items into the four trust dimensions.

#### Building Blocks of Trust Instruments

Complementary instruments for patient-families (*Trust-Building Blocks-Patient-Family*, TRUST-BB-F) and physicians (*Trust-Building Blocks-Physician*, TRUST-BB-MD) were designed for concurrent completion. Review panel deliberation generated new items identified as essential building blocks for forming trust, using a 5-point Likert scale (1=not at all important; 5=extremely important). The same validation approach including expert consultations to align items with individual trust dimensions was applied.

### Phase 2: Instrument Content Validation

All four instruments underwent two validation rounds: (1) dimensional validation to ensure item alignment to the correct trust dimension, and (2) contextual validation to ensure items fit pediatric brief encounter settings.

Validation followed the RAND/UCLA Appropriateness Methodology (RAM), in which expert panels rate item relevance on a 9-point Likert scale (1=not at all relevant; 9=extremely relevant) and provide open feedback. A combined synthetic and human scale-content validity index (S-CVI) threshold of ≥0.80 was set for each instrument.[25,26]

#### Synthetic Instrument Validation (SIV) Framework

The SIV process was governed by three human-centric principles: (1) complementary integration (it augments rather than replaces traditional human expert validation); (2) collaborative framework (human consensus serves as the primary reference standard, synthetic processes being complementary); and (3) human oversight (all SIV-suggested refinements require human expert confirmation and approval).[27]

#### Technical Infrastructure and Model Selection

All selected LLMs and runs were conducted in Perplexity Pro (*Perplexity AI*) workspaces provided with identical context, including the study protocol, RAM booklet,[28] latest instrument iterations, and relevant scoping review.[1] This employment of retrieval-augmented generation (RAG) was strategic to improve the validation accuracy.[29]

Three leading LLMs were selected based on their reasoning abilities (as of September 2025): Claude Sonnet 4[30], Generative Pre-Trained Transformer 5 (GPT-5)[31], and Grok 4[32].

#### Synthetic Expert Development

LLMs were created to embody several expert roles: pediatric emergentologist, pediatric general surgeon, quantitative researcher, emergency department (ED) patient-family representative, and surgical outpatient patient-family representative (for patient-family instruments); and pediatric emergentologist, pediatric general surgeon, and quantitative researcher (for physician instruments). Each synthetic expert was synthetically recruited twice per model to capture varied perspectives within the assigned role.

Prompt design followed best practices using the structure: role details, prompt, instructions, and parameters.[19]

#### Human-Synthetic Collaborative Strategy

A simultaneous parallel evaluation design was conducted between human and synthetic expert panels receiving identical instructions and context. Evaluations occurred simultaneously and independently to ensure human experts remained blind to synthetic outputs.[27] Inter-rater reliability analyses determined comparability of combined human-synthetic panels.

### Two-Phase Content Validation Protocol

#### Dimensional Content Validation

Dimensional validation evaluated each item against its assigned trust dimension (dependability, caring, communication, competence). Three human experts were recruited to evaluate both instrument types. In parallel, Claude Sonnet 4, GPT-5, and Grok 4 models were persona-prompted for patient-family and physician expert roles (**Supplementary Material S2**).

#### Contextual Content Validation

Following dimensional validation, items were evaluated for relevance to pediatric emergency and surgical care contexts. The human panel comprised seven experts for patient-family instruments and five experts for physician instruments. Welch’s t-tests were used to compare I-CVI rankings between each synthetic and human expert panel. Claude Sonnet 4 was selected based on showing lowest variance to human expert panels and was persona-prompted for patient-family and physician expert roles.

### Data analysis

#### Content Validity Index

Item-level (I-CVI) and scale-level (S-CVI) content validity indices were calculated for both validation phases. I-CVI was calculated by dividing the number of experts rating an item as relevant (7-9) by total experts for each instrument, independently for human and synthetic panels and combined. S-CVI was calculated as the average of I-CVIs (S-CVI/Ave) for each instrument, demonstrating the proportion of content-valid items.[14]

#### Comparison of LLM to Human Ratings

Inter-rater reliability was assessed using fixed-marginal Fleiss’ kappa (**κ**), via Justus Randolph’s Fleiss Kappa calculator.[33] Items were categorized as either category one (ratings 0-6) or category two (ratings 7-9) according to RAM interpretation. Values were interpreted as: κ<0.40 (poor), 0.40-0.59 (fair), 0.60-0.79 (substantial), and 0.80-1.00(almost perfect agreement).[34] The target was **κ**≥0.80 for human-synthetic panel agreement.

#### Instrument Refinement

Final expert deliberation determined instrument modifications based on open comments. Significant synthetic panel comments were confirmed with the human panel or survey methodologist expert before implementation.

## RESULTS

### Phase 1: Instrument Design

Following the review of the existing literature, the *Trust in Physician Scale* showed the closest match, and the original author granted full permission for modifications.[23]

The distribution of items across trust dimensions is shown in **Table 1**, with complete instruments provided in **Supplementary Material S3**.

**Table 1.** Distribution of Trust Dimensions Across Four Instruments. Note: TRUST-E-F = Trust-Experience - Patient-Family; TRUST-BB-F = Trust-Building Blocks - Patient-Family; TRUST-P-MD = Trust-Perception - Physician; TRUST-BB-MD = Trust-Building Blocks - Physician.

| Trust Dimension | TRUST-E-F | TRUST-BB-F | TRUST-P-MD | TRUST-BB-MD |
| --- | --- | --- | --- | --- |
| Caring | 2 | 4 | 2 | 4 |
| Competence | 3 | 2 | 2 | 2 |
| Communication | 3 | 2 | 2 | 2 |
| Dependability | 3 | 2 | 2 | 2 |
| General | 1 | 0 | 0 | 0 |
| Total Items | 12 | 10 | 8 | 10 |

### Phase 2: Two-Phase Content Validation

#### Dimensional Content Validation

Three human experts (ED patient-family representative, pediatric emergentologist, quantitative researcher) completed the pilot assessment for both patient-family and physician instruments using the RAM approach.

Thirty synthetic experts completed the assessment for patient-family instruments, with ten experts from each model (Claude Sonnet 4, GPT-5, Grok 4). Eighteen synthetic experts completed the pilot assessment for physician instruments, with 6 experts per model (Claude Sonnet 4, GPT-5, Grok 4). Synthetic patient-family representatives were excluded from the physician validation.

The S-CVI/Ave scores for the human-synthetic expert panel, also shown in **Figure 2**, were: TRUST-E-F 0.98 (SD = 0.02), TRUST-BB-F 0.91 (SD = 0.15), TRUST-P-MD 1.00 (SD = 0), and TRUST-BB-MD 0.91 (SD = 0.17) (I-CVI scores: **Supplementary Material S4**). Each individual panel (Claude Sonnet 4, GPT-5, Grok 4, human) achieved S-CVI ≥0.80 for all instruments.

**Figure 2.**
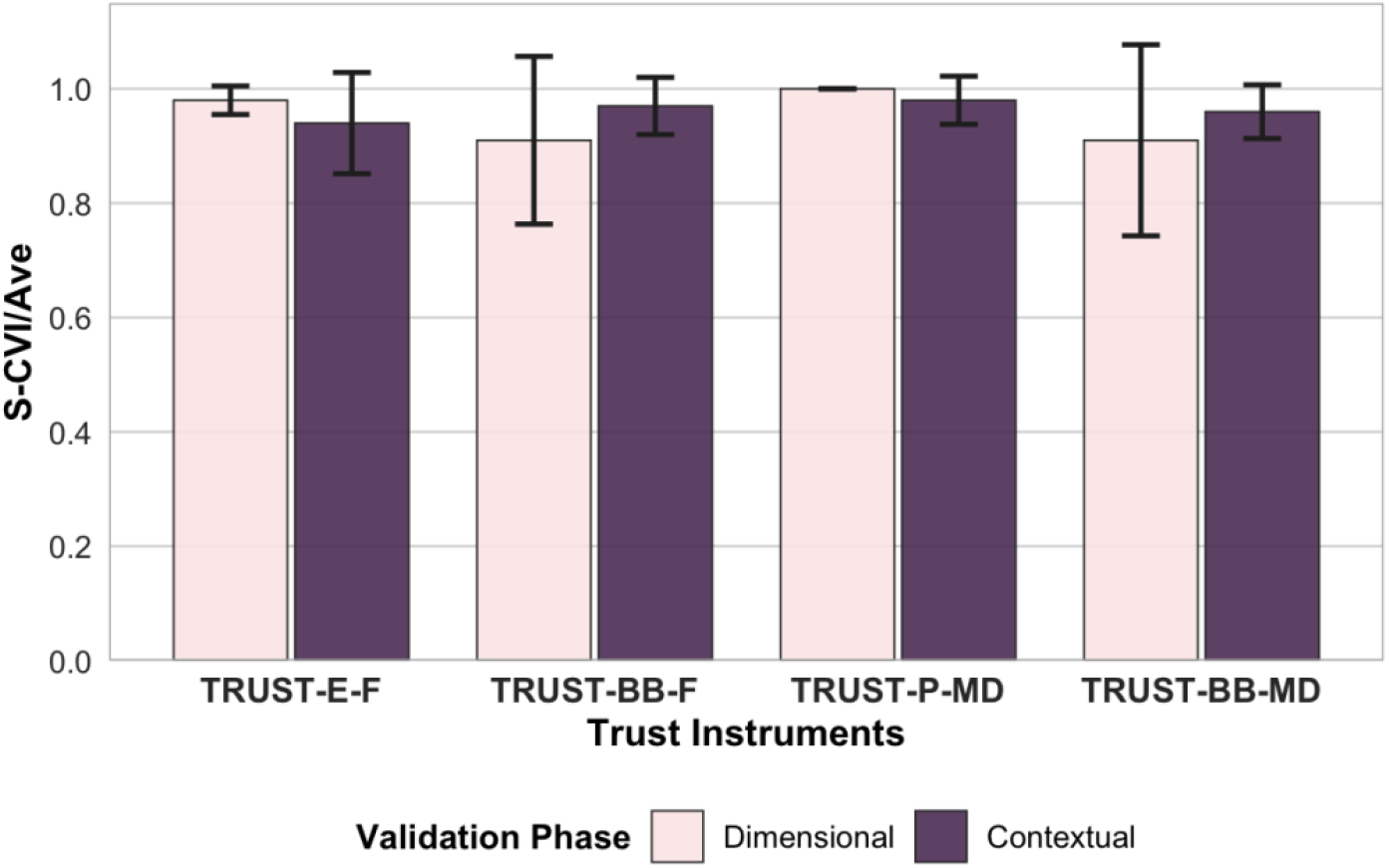
Scale-Content Validity Index (S-CVI/Ave) for trust instruments of the human-synthetic expert panel in contextual and dimensional validation phases. Error bars represent standard deviation of I-CVI scores across items. TRUST-E-F = Trust-Experience - Patient-Family; TRUST-BB-F = Trust-Building Blocks - Patient-Family; TRUST-P-MD = Trust-Perception - Physician; TRUST-BB-MD = Trust-Building Blocks - Physician.

**Figure 3.**
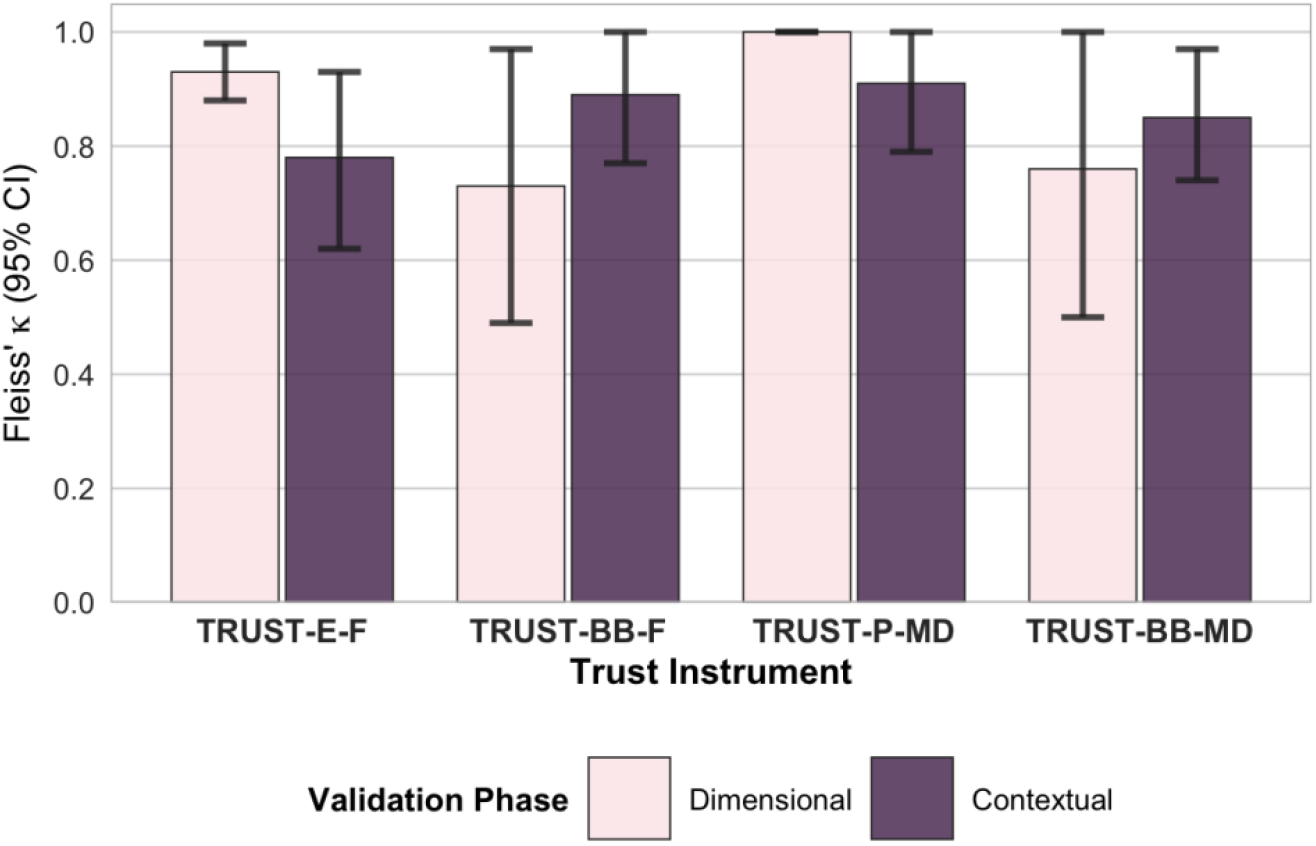
Fleiss’ kappa (k) values of the four trust instruments across dimensional and contextual validation phases. Error bars represent 95% confidence intervals. TRUST-E-F = Trust-Experience - Patient-Family; TRUST-BB-F = Trust-Building Blocks - Patient-Family; TRUST-P-MD = Trust-Perception - Physician; TRUST-BB-MD = Trust-Building Blocks - Physician.

Fixed-marginal kappa values for the combined human-synthetic expert panel were: TRUST-E-F (n = 33; **κ**= 0.93, 95% CI [0.88, 0.98]), TRUST-BB-F (n = 33; **κ**= 0.73,95% CI [0.49, 0.97]), TRUST-P-MD (n = 21; **κ**= 1.00, 95% CI [1.00, 1.00]), TRUST-BB-MD (n=21; **κ**= 0.76, 95% CI [0.50, 1.00]) as shown in **Figure 3**.

Overall instrument agreement was substantial for patient-family instruments (n = 33; **κ**= 0.84, 95% CI [0.72, 0.96]) and physician instruments (n =21; **κ**= 0.87, 95% CI [0.72, 1.00]) as shown in **Figure 4**. The agreement level confirmed acceptable progression to the next validation phase.

**Figure 4.**
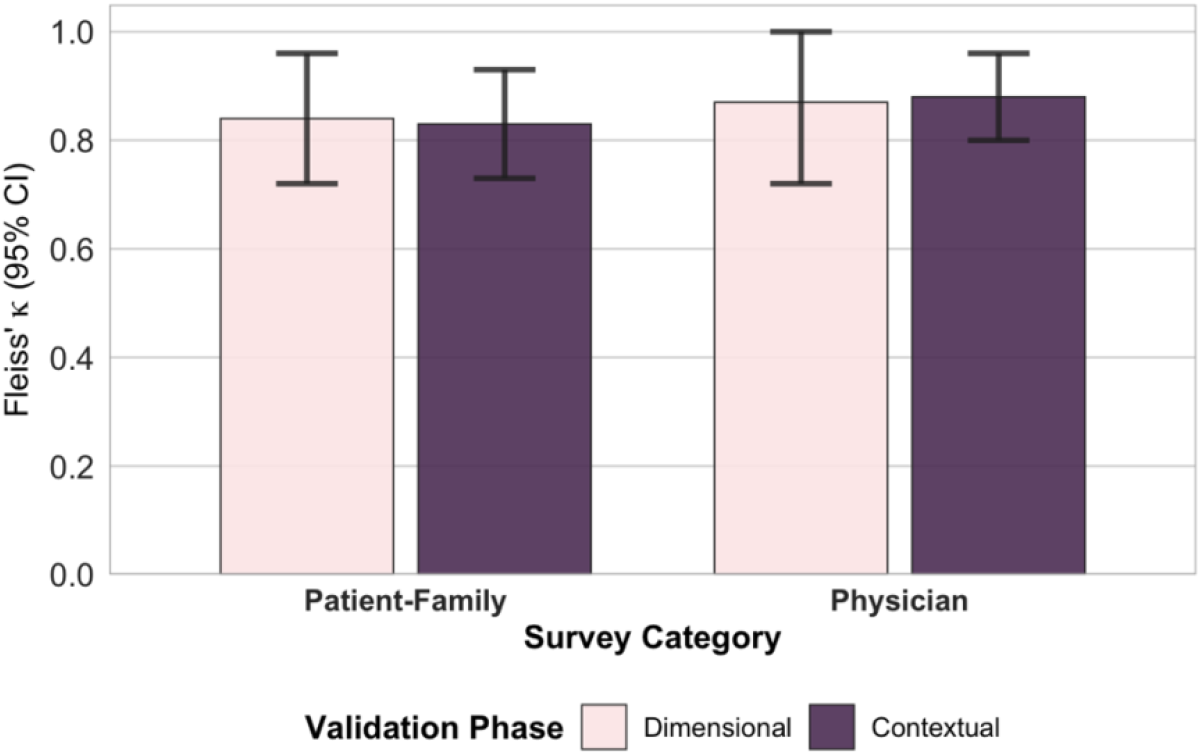
Fleiss’ kappa (**κ**) values of trust instruments per respondent category (patient-family and physician) across dimensional and contextual validation phases. Error bars represent 95% confidence intervals.

#### Contextual Content Validation

Seven human experts (pediatric general surgeon, pediatric emergentologist, pediatric nurse, pediatric occupational therapist, ED patient-family representative, two doctoral students) completed the patient-family instrument assessment, and five human experts (identical except one doctoral student and pediatric nurse) completed the physician instrument assessment. Based on the I-CVI scores of the dimensional content validation, Claude Sonnet 4 showed more non-significant differences from human expert panels (3 of 4 instruments) than GPT-5 or Grok4 (2 of 4 instruments each).

The S-CVI/Ave scores (**Figure 2**) for the human-synthetic expert panel were: TRUST-E-F 0.94 (SD = 0.09), TRUST-BB-F 0.97 (SD = 0.05), TRUST-P-MD 0.98 (SD = 0.04),and TRUST-BB-MD 0.96 (SD = 0.05) (I-CVI scores: **Supplementary Material S4**). Both Claude Sonnet 4 and human panels achieved S-CVI ≥0.80 for all instruments. Fixed-marginal kappa values for the combined human-synthetic expert panel were: TRUST-E-F (n = 17; **κ**= 0.78, 95% CI [0.62, 0.93]), TRUST-BB-F (n = 17; **κ**= 0.89,95% CI [0.77, 1.00]), TRUST-P-MD (n = 11; **κ**= 0.91, 95% CI [0.79, 1.00]), TRUST-BB-MD (n = 11; **κ**= 0.85, 95% CI [0.74, 0.97]) as shown in **Figure 3**.

Overall instrument agreement was: patient-family instruments (n=17; **κ**= 0.83, 95% CI [0.73, 0.93]) and physician instruments (n = 11; **κ**= 0.88, 95% CI [0.80, 0.96]) as shown in **Figure 4**.

#### Instrument Refinement

Refinements included adjusting words for better contextual fit, removing double-barreled aspects where items covered two or more concepts at once, and bolding aspects of the item that attract respondents to the main point (**Supplementary Material S3**).[35,36] One TRUST-E-F item (“I felt my child’s doctor was honest with me about my child’s condition”) was removed based on human expert majority (4 of 7) despite favorable I-CVI scores (dimensional: 0.97, contextual: 0.94) due to apparent redundancy with item 3, which received perfect I-CVI in both phases.

## DISCUSSION

This study content validated four pediatric trust instruments (TRUST-E-F, TRUST-BB-F, TRUST-P-MD, TRUST-BB-MD) using parallel human and synthetic expert panels through a new approach termed SIV.

All instruments exceeded the established S-CVI/Ave threshold of 0.80, by achieving scores above 0.90 in both dimensional and contextual validation phases. These results surpass the content validity literature benchmarks of panels greater than 5 experts, and suggest that instruments appropriately measure trust dimensions while maintaining contextual relevancy to pediatric emergency and surgical care settings.[25] Inter-rater reliability among human and synthetic experts demonstrated substantial to almost perfect agreement. Notably, even the lowest-performing individual instrument (TRUST-BB-F, **κ**= 0.73) approached the substantial agreement threshold, while aggregate patient-family (**κ**= 0.83-0.84) and physician (**κ**= 0.87-0.88) instruments consistently surpassed the 0.80 target. This consistency across diverse experts demonstrates that LLMs can replicate similar evaluations to human expert panels.

Traditional content validation typically requires panels of five to ten experts over extended timelines.[25] Our SIV approach completed both validation phases within four weeks with a minimum of 11 experts evaluating the instrument and maintaining robust validity metrics. This addresses fundamental barriers in specialized populations where expert availability is limited such as pediatrics.[16,37] This acceleration is aligned with emerging evidence that LLMs can achieve strong agreement with human evaluators in healthcare contexts. For example, GPT-o3-mini achieved an intraclass correlation coefficient of 0.82 with human expert evaluators when evaluating electronic health record quality, while GPT-4o has shown validity as a psychological text classifier.[19,38] Yet, most AI applications have focused on evaluating completed instruments rather than developing new ones, making this study among the first to demonstrate LLMs potential in preliminary content validation phases.[27]

Persona-prompted LLMs successfully embodied specific expert roles, which is highlighted by role-appropriate reasoning in the open comments. For example, a synthetic patient-family representative (Claude Sonnet 4, dimensional phase) noted: “As a parent, my confidence in the doctor’s ability to handle my child’s medical issues is fundamental to trust.” When combined with RAG, persona prompting can improve judgment accuracy by 30%, which supports the approach’s effectiveness for subjective, context-dependent validation tasks.[29]

A key pattern emerged where synthetic experts demonstrated tendencies to rate items more positively while human experts provided more critical evaluations. This complementarity dynamic of “synthetic optimism” and “human conservatism” was advantageous; synthetic optimism ensured efficient consensus-building and progression through validation stages, while human skepticism identified opportunities for refinement.[39] This suggests that human and synthetic experts may function synergistically rather than as interchangeable panels, aligning with the collaborative AI frameworks where differing capabilities are meant to augment rather than replace one another.[40]

The SIV framework has the potential for extension to subsequent validation phases including cognitive interviewing and comprehensive psychometric assessment.[15,26,41] Once the potential for SIV to function within the entire instrument validation process, it should be evaluated for its applicability to broader healthcare contexts such as oncology, mental health and quality of care measurement.[42] However, adaption would require context specific persona development and equally rigorous validation of synthetic response patterns against real populations.[43] Standardized guidelines for SIV implementation include optimal LLM selection criteria, human-synthetic panel composition and the need to establish quality assessment. As AI is involved to a greater extent within instrument validation, it is important to ensure transparency and accountability of its use by integrating frameworks such as FUTURE-AI.[44]

## Limitations

Our study has several limitations. First, the rapid advancement of LLM capabilities means that the models evaluated in this study (up to September 2025) may not reflect state of the art performance by the time of publication. Second, this study focused exclusively on content validation. The applicability and validity of SIV for subsequent validation phases such as cognitive interviewing, psychometric property assessment, and pilot testing remains to be empirically established. Third, the study was conducted within a single healthcare context, which may limit its generalizability, requiring evaluations to establish the robustness of SIV across diverse settings and constructs. Additionally, while inter-rater reliability between human and synthetic experts was substantial, systematic differences in rating patterns, i.e., “synthetic optimism” and “human conservatism”, suggest that these expert types may not be fully interchangeable. Understanding the sources of these differences and their implications for validation requires further investigation. Finally, the study relied on validation metrics designed for human-only expert panels; modified thresholds or alternative metrics for hybrid panels merit exploration.

## Conclusion

This study demonstrates the feasibility and validity of using the SIV approach with persona-prompted LLMs to augment content validation of healthcare instruments. Our four pediatric trust instruments successfully achieved validation thresholds with substantial human-synthetic expert agreement, enabling their progression to psychometric testing. Beyond validating these specific instruments, this work establishes a methodological blueprint for accelerating instrument development.[2]

The SIV approach addresses critical barriers that have historically constrained healthcare instrument development.[45,46] As LLM capabilities continue to evolve, AI-assisted validation offers a scalable pathway to accelerate instrument development in healthcare contexts where timely, valid measurement tools are critically needed to advance evidence-based patient care.

## Data Availability

The authors confirm the data supporting this study may be found within the manuscript and its Supplementary Material. Raw data may be requested from the corresponding author, upon reasonable request.

## ACKNOWLEDGEMENTS

The authors would like to express their sincere gratitude to the participating physicians, nurses, health services researchers and patient family representatives for their valuable contributions to this study. The authors would also like to thank Professor Jeannie Haggerty for providing foundational background to instrument design. Three large language models (Claude Sonnet 4 (Anthropic, 2025), GPT-5 (OpenAI, 2025), and Grok4 (xAI, 2025)) were used as synthetic expert panels for instrument validation. Besides this major use of LLMs as part of the described Methods, Claude Sonnet 4 & Claude Sonnet 4.5 (Anthropic, 2025) were used through PerplexityPro to improve manuscript readability, including checking grammar, clarity, and flow of text, and for figure code debugging. The author(s) reviewed and edited all AI-generated content and take(s) full responsibility for all manuscript content.

## AUTHOR CONTRIBUTIONS

Ella Boone, Katya Loban and Dan Poenaru (Conceptualization), All Authors (Methodology), Ella Boone (Data Curation, Formal Analysis, Writing - Original Draft), All Authors (Writing - Review and Editing), Dan Poenaru (Supervision)

