## Supplementary Material for "Synthetic Validation of Pediatric Trust Instruments using Persona-Driven Large Language Models"

**SUPPLEMENTARY DATA**

**S1 Wake Forest Physician Trust Scale Based Item Generation**

**Table S1.** Adaption of Wake Forest Physician Trust Scale to develop the TRUST-E-F (Trust- Experience- Patient-Family) instrument. Bolded items were adapted from the original instrument and contextualized for pediatric emergency and surgical care settings.

| WFPTS Items | TRUST-E-F Items |
| --- | --- |
| *Response categories: Strongly Agree, Agree, Neutral, Disagree, Strongly Disagree*   1. **[Your doctor] will do whatever it takes to get you all the care you need**. 2. Sometimes [your doctor] cares more about what is convenient for [him or her] than about your medical needs. 3. **[Your doctor’s] medical skills are not as good as they should be.** 4. **[Your doctor] is extremely thorough and careful.** 5. **You completely trust [your doctor’s] decisions about which medical treatments are best for you.** 6. **[Your doctor] is totally honest in telling you about all of the different treatment options available for your condition.** 7. **[Your doctor] only thinks about what is best for you.** 8. **Sometimes [your doctor] does not pay full attention to what you are trying to tell [him or her].** 9. You have no worries about putting your life in [your doctor’s] hands. 10. **All in all, you have complete trust in [your doctor].** | *Response categories: 1= No, not at all 2= No, not really 3= Yes, a little 4= Yes, definitely*   1. **I felt my child’s doctor did whatever it took to get my child all the care they needed.** 2. My child's doctor made me feel that they genuinely cared about my child's health. 3. **I felt my child's doctor could manage the specific medical problems we were seeking care for.** 4. **My child’s doctor appeared extremely thorough in their consultation.** 5. **I trusted the decisions my child's doctor made about my child's medical care.** 6. **I felt my child’s doctor was honest with me about my child’s condition.** 7. **I believe my child’s doctor only thought about what was best for my child.** 8. **My child’s doctor seemed to pay full attention to what we are trying to tell them, or my child was trying to tell them.** 9. My child's doctor seemed to be open and transparent when discussing my child’s condition, even when the news was challenging. 10. I felt as though my child's doctor focused on what was best for my child’s medical needs. 11. My child's doctor explained the risks and benefits of treatments clearly to me.   Standalone question:  *Response categories: 1= Not at all 2= A little 3= Moderately 3= A lot 4= A great deal*   12. **Overall, how much do you trust your child's doctor?** |
| *Modifications due to expert deliberation (Item order of ETP is rearranged to mirror WFPTS for ease of comparison, unbolded items are newly designed items):*  For all items: Changed “your doctor” to “my child’s doctor” and all other indications about adult primary care to “my child”, response category changed to a 4-point Likert scale to remove neutral option, verb tense changed to past due to acute care setting  Item 1: Added “I felt” to highlight the patient-family/caregiver reflection  Item 3: Added “I felt” to highlight the patient-family/caregiver reflection, positive wording of competence  Item 4: Added “appeared” to highlight that is how the patient-family/caregiver is feeling, as they may not know general standards of care for what they are seeking  Item 5: Removed completely due to brief nature of pediatric setting and lack of prior relationship  Item 6: Removed “totally” to highlight patient-family/caregiver reflection that may not know or have enough time to be aware of their child’s condition and generalized from treatments to condition to condition to highlight nature of surgical and emergency consultation  Item 7: I believe is added to highlight patient-family/caregiver opinion  Item 8: Added layer of communication to both child and patient-family/caregiver, as well as reflection of their opinion  Item 10 (12 -ETP): “all in all” to “overall” to highlight entire consultation experience, given own Likert scale quantify the experience of trust from the one visit | |

**S2 Sample Synthetic Dimensional Prompt**

All expert prompts followed the same outline with differences in Role and Role Details, respondent group (physician or patient-family), as well as the instrument that was included, all other components were identical.

**Table S2.** Sample prompt for the Trust- Experience- Patient-Family instrument validation using Perplexity models.

| ***Prompt Component*** | Sample Description |
| --- | --- |
| ***Role*** | Pediatric Surgeon |
| ***Role Details***  *What context is the synthetic data produced?* | Pretend you are a pediatric general surgeon, working primarily at an academic children’s hospital. |
| ***Prompt***  *What’s the study context and goals?* | You are asked to evaluate the content validity of a survey designed to understand how and what forms trust in pediatric surgical and urgent care settings. This survey is designed for parents/guardians of patients who are waiting to receive urgent care or after they had a surgical consultation. The files attached describe the study that will be conducted at the Montreal children’s hospital and the systematic review used to identify the need for this work and design our working definition of trust and its dimensions (competency, dependability, caring, communication).  We require your input in ranking the relevance of each item in this survey to the dimension of trust it is measuring of either competency, dependability, caring and communication. I will ask you to do this by rating each of the numbered items on a scale of 1 to 9 where 1 is not at all relevant and 9 is extremely relevant. |
| ***Instructions***  *What data should the LLM synthetically provide you?* | Please review the documents and read the instructions and scale given to future respondents- *Instructions: Based on your experience during today’s visit or during your most recent visit in a paediatric surgical or emergency setting, please rate how much you agree with each statement about your child’s doctor. Scale 1= No, not at all 2= No, not really 3= Yes, a little 4= Yes, definitely.*  Based on the dimension of trust next to each item; rate the relevance of each item INDIVIDUALLY to that dimension scale of 1 (not at all relevant) to 9 (extremely):   I. My child's doctor made me feel that they genuinely cared about my child's health. - CARING II. I felt my child's doctor could manage the specific medical problems we were seeking care for. -COMPETENCE III. My child's doctor seemed to be open and transparent when discussing my child’s condition, even when the news was challenging. -COMMUNICATION IV. I trusted the decisions my child's doctor made about my child's medical care. - COMPETENCE V. I felt as though my child's doctor focused on what was best for my child’s medical needs. -CARING VI. My child's doctor explained the risks and benefits of treatments clearly to me. - COMMUNICATION VII. I felt my child’s doctor was honest with me about my child’s condition. - DEPENDABILITY VIII. I believe my child’s doctor only thought about what was best for my child. - DEPENDABILITY IX. My child’s doctor paid full attention to what we are trying to tell them, or my child was trying to tell them. - COMMUNICATION X. My child’s doctor appeared extremely thorough in their consultation. – COMPETENCE XI. I felt my child’s doctor did whatever it took to get my child all the care they needed. - DEPENDABILITY   There is a standalone question with it’s own scale (1= Not at all 2= A little 3= Moderately 3= A lot 4= A great deal) but the same instructions, please rank the following item on the same scale of 1 to 9:   I. Overall, how much do you trust your child’s doctor? -OTHER |
| ***Parameter***  *How should the data be displayed?* | Please provide your answer to each individual item in a table format with a column for: the item number, the item you are evaluating, your rating of the item, any clarification or comments about the wording, syntax, appropriateness |
| ***Files***  *What will be given to provide identical context to human experts?* | Approved study protocol (Jul 10, 2025), prior systematic review, physician and patient-family instrument document, RAND/UCLA appropriateness methods protocol |

**S3 Initial and Final Trust Instrument Iterations**

**Table S3.** Instruments before and after content validation

*Note: TRUST-E-F = Trust-Experience - Patient-Family; TRUST-BB-F = Trust-Building Blocks - Patient-Family; TRUST-P-MD = Trust-Perception - Physician; TRUST-BB-MD = Trust-Building Blocks - Physician.*

| **TRUST-E-F** | |
| --- | --- |
| *Initial* | *Final* |
| *Instructions: Based on your experience during today’s visit or during your most recent visit in a paediatric surgical or emergency setting, please rate how much you agree with each statement about your child’s doctor.*  *Response categories: 1= No, not at all 2= No, not really 3= Yes, a little 4= Yes, definitely*   1. My child's doctor made me feel that they genuinely cared about my child's health. 2. I felt my child's doctor could manage the specific medical problems we were seeking care for. 3. My child's doctor seemed to be open and transparent when discussing my child’s condition, even when the news was challenging. 4. I trusted the decisions my child's doctor made about my child's medical care. 5. I felt as though my child's doctor focused on what was best for my child’s medical needs. 6. My child's doctor explained the risks and benefits of treatments clearly to me. 7. I felt my child’s doctor was honest with me about my child’s condition. 8. I believe my child’s doctor only thought about what was best for my child. 9. My child’s doctor seemed to pay full attention to what we are trying to tell them, or my child was trying to tell them. 10. My child’s doctor appeared extremely thorough in their consultation. 11. I felt my child’s doctor did whatever it took to get my child all the care they needed.     Standalone question:  12. Overall, how much do you trust your child's doctor?  *Response categories: 1= Not at all 2= A little 3= Moderately 3= A lot 4= A great deal* | *Instructions: Based on your experience during today’s visit or during your most recent visit in a paediatric surgical or emergency setting, please rate how much you agree with each statement about your child’s doctor.*  *Response categories: 1= No, not at all 2= No, not really 3= Yes, a little 4= Yes, definitely*   1. My child's doctor made me feel that they **genuinely cared** about my child's health. 2. I felt my **child's doctor could** **manage** the specific medical problems we were seeking care for. 3. My child's doctor seemed **transparent** **when discussing** my child’s condition, even when the news was challenging. 4. I had **confidence in the** **recommendations** my child's doctor made about my child's care. 5. I felt as though my child's doctor **focused on what was best** for my child’s medical needs. 6. My child's doctor **explained the outcomes of treatments clearly** to me. 7. I believe my child’s doctor made **decisions that are in the best interest** of my child’s care 8. My child’s doctor seemed **to pay full attention** to what we are trying to tell them, or my child was trying to tell them. 9. I felt my child’s doctor was **extremely thorough** in their consultation. 10. I felt my child’s doctor **did whatever it took** to get my child all the care they needed.     Standalone question:   11. Overall, how much do you **trust** your child's doctor?     *Response categories: 1= Not at all 2= A little 3= Moderately 3= A lot 4= A great deal* |
| **TRUST-BB-F** | |
| *Instructions: Please think about some of the important ingredients in building and maintaining trust in your child's doctor across all stages of care in paediatric surgical or emergency settings. Please rate the following:*  *Response categories: 1 = Not at all important, 2 = Slightly important, 3 = Moderately important, 4 = Very important, 5 = Extremely important*   1. My child’s doctor explaining and answering all my questions and concerns about my child’s condition and treatment. 2. My child’s doctor managing my child’s pain effectively throughout the treatment. 3. My child’s doctor speaking directly to my child (when age appropriate), making sure my child is also included in the conversation. 4. My child’s doctor communicating clearly about what is known and not known about my child’s condition or treatment. 5. My child’s doctor respecting my role as a parent/guardian. 6. My child’s doctor following through on what they said they would do during our visit. 7. My child’s doctor showing genuine concern for my child’s well-being across all stages of care. 8. My child’s doctor treating our family with respect across all stages of care. 9. My child’s doctor making an effort to understand our concerns during our visit. 10. My child’s doctor spending the appropriate amount of time with us during our visit. | *Instructions: Please think about some of the important ingredients in building and maintaining trust in your child's doctor across all stages of care in paediatric surgical or emergency settings. Please rate the following:*  *Response categories: 1 = Not at all important, 2 = Slightly important, 3 = Moderately important, 4 = Very important, 5 = Extremely important*   1. My child’s doctor **knowledgeably addressing** my concerns about my child’s treatment. 2. My child’s doctor **managing** my child’s treatment effectively. 3. My child’s doctor **speaking directly** to my child (when age appropriate), making sure my child is also included in the conversation. 4. My child’s **doctor communicating clearly** about the uncertainties of my child’s condition or treatment. 5. My child’s doctor **consistently** respecting my input as a parent/guardian. 6. My child’s doctor **following through** on what they said they would do during our visit. 7. My child’s doctor showing **genuine concern** for my child’s well-being across all stages of care. 8. My child’s doctor treating our family with **respect** across all stages of care. 9. My child’s doctor **making an effort** to understand our perspective during our visit. 10. My child’s doctor spending the **appropriate amount** of time with us during our visit. |
| **TRUST-P-MD** | |
| *Instructions: Based on your experience during your most recent shift in a paediatric surgical or emergency setting, please rate how much you believe patient-families and patients trusted you in each of the following aspects of care.*  *Response categories: 1= No, not at all 2= No, not really 3= Yes, a little 4= Yes, definitely*   1. My clinical expertise and knowledge. 2. My ability to make the right decisions for a child’s care. 3. My honesty and openness when sharing information. 4. My ability to explain things clearly. 5. My reliability in following through on what I say. 6. My commitment to acting in the child’s best interest. 7. My respect for each family’s background and values. 8. My compassion and empathy toward patients and families. | *Instructions: Based on your experience during your most recent shift in a paediatric surgical or emergency setting, please rate how much you believe patient-families and patients trusted you in each of the following aspects of care.*  *Response categories: 1= No, not at all 2= No, not really 3= Yes, a little 4= Yes, definitely*   1. My clinical **expertise** relevant to the child’s condition. 2. My ability to make the **right decisions** for a child’s care. 3. My honesty and **openness** when sharing information. 4. My ability to explain things **clearly**. 5. My **reliability** in following through on what I say. 6. My **commitment** to acting in their best interest. 7. My **respect** for each family’s background and values. 8. My **compassion** toward patients and families. |
| **TRUST-BB-MD** | |
| *Instructions: In your experience, how important do you believe each of the following factors is in developing parents'/patients' trust in you as a healthcare provider across all stages of care you are involved in?*  *Response categories: 1 = Not at all important, 2 = Slightly important, 3 = Moderately important, 4 = Very important, 5 = Extremely important*   1. Showing concern for a child’s comfort and overall well-being. 2. Explaining and answering questions and concerns about care from patient-families sufficiently. 3. Speaking directly to the child (when age-appropriate) and including them in the conversation. 4. Involving parents in decision about their child’s care. 5. Effectively managing child’s pain and symptoms. 6. Being honest about what is known and not known regarding a child’s condition or treatment. 7. Treating patient-families with respect. 8. Ensuring patient-families feel understood in their concerns. 9. Following through on actions and information discussed during their visit. 10. Spending enough time with patient-families during their visit. | *Instructions: In your experience, how important do you believe each of the following factors is in developing parents'/patients' trust in you as a healthcare provider across all stages of care you are involved in?*  *Response categories: 1 = Not at all important, 2 = Slightly important, 3 = Moderately important, 4 = Very important, 5 = Extremely important*   1. Showing **concern** for a child’s comfort and overall well-being. 2. Conveying my **clinical expertise** when addressing patient-family concerns 3. **Speaking directly** to the child (when age-appropriate), including them in the conversation. 4. **Consistently** involving parents in decisions about their child’s care. 5. **Effectively** managing child’s treatment across all stages of care. 6. **Being honest** about what is known and not known regarding a child’s condition or treatment. 7. Treating patient-families with **respect**. 8. Ensuring patient-families **feel understood** in their concerns. 9. **Following through** on actions and information discussed during their visit 10. **Not appearing rushed** during consultations with patient-families during their visit. |

**S4 Item-Content Validity Index Scores - Dimensional and Contextual Content Validation Phases**

**Table S4.** I-CVI values for the human-synthetic panel across the trust instruments.

*Note: TRUST-E-F = Trust-Experience - Patient-Family; TRUST-BB-F = Trust-Building Blocks - Patient-Family; TRUST-P-MD = Trust-Perception - Physician; TRUST-BB-MD = Trust-Building Blocks - Physician.*

| **Instrument** | **Item #** | **Dimensional** | **Contextual** |
| --- | --- | --- | --- |
| **TRUST-E-F**  **(n = 33, 17)** | **1** | 0.97 | 0.94 |
|  | **2** | 1.00 | 1.00 |
|  | **3** | 1.00 | 1.00 |
|  | **4** | 0.94 | 0.82 |
|  | **5** | 1.00 | 1.00 |
|  | **6** | 1.00 | 1.00 |
|  | **7** | 0.97 | 0.94 |
|  | **8** | 1.00 | 0.71 |
|  | **9** | 1.00 | 0.94 |
|  | **10** | 0.97 | 0.94 |
|  | **11** | 1.00 | 1.00 |
|  | **12** | 0.94 | 0.94 |
| **TRUST-BB-F**  **(n = 33, 17)** | **1** | 0.64 | 1.00 |
|  | **2** | 0.97 | 0.88 |
|  | **3** | 0.97 | 1.00 |
|  | **4** | 1.00 | 1.00 |
|  | **5** | 0.64 | 0.94 |
|  | **6** | 1.00 | 1.00 |
|  | **7** | 0.97 | 1.00 |
|  | **8** | 1.00 | 1.00 |
|  | **9** | 1.00 | 1.00 |
|  | **10** | 0.88 | 0.88 |
| **TRUST-P-MD**  **(n = 21, 11)** | **1** | 1.00 | 1.00 |
|  | **2** | 1.00 | 1.00 |
|  | **3** | 1.00 | 1.00 |
|  | **4** | 1.00 | 0.91 |
|  | **5** | 1.00 | 1.00 |
|  | **6** | 1.00 | 1.00 |
|  | **7** | 1.00 | 0.91 |
|  | **8** | 1.00 | 1.00 |
| **TRUST-BB-MD**  **(n = 21, 11)** | **1** | 1.00 | 1.00 |
|  | **2** | 0.57 | 0.91 |
|  | **3** | 1.00 | 0.91 |
|  | **4** | 0.62 | 1.00 |
|  | **5** | 0.95 | 0.91 |
|  | **6** | 1.00 | 1.00 |
|  | **7** | 1.00 | 1.00 |
|  | **8** | 1.00 | 1.00 |
|  | **9** | 1.00 | 1.00 |
|  | **10** | 0.95 | 0.91 |
